## Supplementary material for "Autistic adults’ experiences of accessing and receiving mental health care and their priorities for improvements: a qualitative study": Topic Guide

Thank you for agreeing to take part in this interview about autistic adults’ views on priorities for improving mental health care. The interview should last about an hour.

*Ethical reminders*

- Remind the person that they have already given consent, and check that they are still ok with this.

- Check the participant is ok with you recording the interview.

- Confirm that you will start recording

For this interview, I will be asking you questions about your views on priorities for improving mental health care for autistic people. We won’t be directly exploring your own experiences of mental health difficulties, only in your perspectives and experiences of accessing and receiving care and your priorities for its improvement. There are no right or wrong answers, please feel free to say as little or as much as you like. We’ll anonymise the information you give us, so we’ll remove anything that could identify you when we type up the interview. Please do let me know if you need to take a break or stop.

**Questions:**

**Part 1: Accessing care**

1. What made you realise you required professional support for your mental health?

*What condition/mental health problem?*

*How did you feel when you realised?*

*(If relevant) what might have prompted you to seek help earlier?*

2. What has your experience of trying to get mental health treatment been like?

*How easy/difficult was it?*

*Were there any barriers to you receiving the support you required? If so, what were they?*

***How could it be improved?***

3. How confident do you feel in accessing mental health support when you need it now?

*Do you know who to contact?*

*Are they easily contactable?*

*How confident are you that you will get a positive response?*

*(If relevant) How could this be improved?*

**Part 2: Experience of mental health care**

1. Can you tell me about the mental health care that you receive or have received?

*What form of care did you receive?*

*In what kind of setting(s) (unclear) have you received care? List of settings in case they’re unaware*

*How did/do you feel in those settings?*

*Did/do you get the support that you need?*

*Did/does anything make it difficult to get the support that you needed/need? If so, what?*

2. How well suited to your needs is/was the treatment you are receiving/received as an autistic person?

*What, if anything, did you find difficult about the treatment?*

*What would have made it easier for you?*

3. What has your experience been like with communicating with mental health professionals?

*How confident did/do you feel explaining your concerns to them?*

*How knowledgeable do you feel mental health professionals are about autism?*

*How well do you think you are understood by mental health professionals?*

*How well are you able to understand mental health professionals?*

*(If relevant) What could mental health professionals do differently to help your understanding?*

*Pharmacological treatments – were they explained well enough? (side effects)*

4. How do you feel about the environment in which you receive(ed) care?

*Was/is there anything you find uncomfortable/distressing? If so, what?*

*Was/is there anything that makes you feel more at ease? If so, what?*

*(If relevant) How could mental health environments be adapted to suit your needs?*

**Part 3: Facilitators**

1. What, if anything, do you do to make your experience of receiving mental health care easier?

*Do you have any coping mechanisms that make your experience easier?*

*Is there anything that you would like to be able to do that would make your experience easier?*

2. What, if anything, could mental health professionals do to make your experience of mental health services easier?

*What do you think they could do more/less of?*

3. What would you like mental health professionals to know about you before working with you?

e.g. needing a break

*What could they do to help make the experience a positive one?*

*Is there anything they could say to make you feel more comfortable?*

*Is there anything that you would not like them to say?*

**Part 4: Priorities for improving care**

1. What are the most important changes that you would like to see that would improve mental health care for autistic people?

*What could staff do before your appointment?*

*What could staff do during your appointment?*

*What could staff do after your appointment?*

*Physical environment?*

*How could treatment be adapted?*

*Which would make the most difference in terms of improving mental health care for you?*

*Anything else you’d like to add/ask?*
