## Supplementary material for "Autistic adults’ experiences of accessing and receiving mental health care and their priorities for improvements: a qualitative study": Interview Debriefing Sheet

**Appendix B**

**Interview Debriefing Sheet**

UCL Research Ethics Committee Approval ID Number: 25209/001

**Title of study:** Autistic adults’ views on priorities for improving mental health care in the UK: a qualitative interview study

We would like to take this opportunity to **thank you** for taking part in this interview. The information you have given us is very valuable and we appreciate your time.

We hope that taking part in the interview has been interesting, although we also appreciate that talking about personal experiences can be difficult.

If you require further emotional support following this interview you might want to:

- Talk with a relative, friend or other supporter if that is what you prefer
- Contact your GP, or a mental health professional if you are currently using services
- Use a publicly available source of emotional support, we have included a list of some options below:

Saneline

They provide emotional support and information: 0300 304 7000. Open 4.30pm – 10.30pm every day. <http://www.sane.org.uk/what_we_do/support/helpline>

Samaritans

Call free any time, from any phone, on 116 123

Contact a Samaritan: If you need someone to talk to, we listen & won't judge or tell you what to do. <https://www.samaritans.org/how-we-can-help/contact-samaritan/>

The Mix

The Mix is a UK based charity that provides free, confidential support for young people under 25 via online, social and mobile.

<https://www.themix.org.uk/>

0808 808 4994

MIND Information and Support

<https://www.mind.org.uk/information-support/>

MIND Infoline

0300 123 3393

Hub of Hope

The Hub of Hope is the UK’s leading mental health support database. It is provided by national mental health charity, Chasing the Stigma, and brings local, national, peer, community, charity, private and NHS mental health support and services together in one place.

<https://hubofhope.co.uk/>

Online Mental Health Support Forum

<http://www.sane.org.uk/what_we_do/support/supportforum/support_rooms/>

CalmZone

Campaign Against Living Miserably

<https://www.thecalmzone.net/help/get-help/>

If you would like to contact a member of the research team at a later date about any queries or concerns that you may have about your taking part in this research, please do not hesitate to contact:

| NAMES | Frederick Taylor (Study Researcher); or  Professor Sonia Johnson (Principal Investigator) |
| --- | --- |
| ADDRESS | Division of Psychiatry, 6^th^ Floor, Maple House, 149 Tottenham Court Road, W1T 7NF |
| EMAIL |  |

If you have given your consent for us to contact you with the findings of this study, these will be sent to you via the contact details you have given us within six months of the study ending.

**Thank you so much again for taking part in our research study.**
